# Premature white matter microstructure in female children with a history of concussion

**DOI:** 10.1101/2021.12.02.21267220

**Authors:** Eman Nishat, Sonja Stojanovski, Shannon E Scratch, Stephanie H Ameis, Anne L Wheeler

## Abstract

As maturation of the brain continues throughout development, there is a risk of interference from concussions which are common in childhood. A concussion can cause widespread disruption to axons and inflammation in the brain and may influence emerging cognitive abilities. Females are more likely to experience persistent problems after a concussion, yet the sex-specific impact of concussions on brain microstructure in childhood is not well understood.

In children from a large population sample, this study (1) investigated differences in white matter and cortical microstructure between children with and without a history of concussion, and (2) examined relationships between altered brain microstructure and cognitive performance.

Neurite density measures from diffusion weighted magnetic resonance imaging were examined in 9-to 10-year-old children in the Adolescent Brain Cognitive Development Study with (*n* = 336) and without (*n* = 7368) a history of concussion. (1) Multivariate regression models were used to investigate the relationships between concussion history, sex, and age in the deep white matter, superficial white matter, subcortical structures, and cortex. (2) Principal component analysis was performed on neurite density, and components were examined in relation to performance on the Flanker Inhibitory Control and Attention Task and the Pattern Comparison Processing Speed Task to investigate the relationship between altered neurite density and cognitive performance.

Neurite density in all tissue types demonstrated robust positive relationships with age reflecting maturation of brain microstructure. (1) Comparisons between children with and without a history of concussion revealed higher neurite density in deep and superficial white matter in females with concussion. No group differences were observed in subcortical or cortical neurite density. (2) Higher neurite density in superficial white matter beneath the frontal and temporal cortices was associated with lower scores on the processing speed test in females with concussion, and higher scores on the processing speed test in males with concussion.

These findings suggest that concussion in childhood leads to premature white matter maturation in females and that this may be associated with slower processing speed. These sex-specific effects on the developing brain may contribute to the enhanced vulnerability to persistent symptoms after concussion in females.

## Introduction

Two-thirds of all concussions occur in children, with 20-40% reporting at least one concussion by the time they reach adolescence.^1–3^ Concussion symptoms include cognitive impairment, prominently problems with attention and processing speed,^4–7^ along with emotional and physical problems, and sleep dysregulation all of which can impact academic performance and overall quality of life.^8–10^ Most children that experience a concussion recover within two weeks of injury, however, 10-30% of children experience symptoms that last longer than 3 months,^11–13^ with females at greater risk of persistent symptoms.^14–16^ Little is known about the association between sex-specific effects of concussion on the developing brain and emerging cognitive abilities.

Brain microstructure continues to mature throughout childhood. Prominently, in both white and gray matter, axons increase in diameter and continue to myelinate, and in white matter bundles, axons also become more tightly packed together.^17–19^ The maturation of these structures is crucial for efficient and coordinated communication in the brain that enables higher order cognitive function.^20^ In recent decades, neuronal changes during brain development have been further described *in vivo* with diffusion MRI methods that infer microstructural properties of brain tissues through the movement of water molecules.^21–23^ These studies have observed earlier maturation of white matter microstructure in females.^24^ The majority of these studies have examined fractional anisotropy (FA) from diffusion tensor imaging (DTI), however, recent studies suggest that measures of neurite density (ND) from more advanced multi-compartment models of water diffusion are more sensitive to the maturation of brain microstructure.^25–28^ ND derived from cylindrically restricted intracellular diffusion is believed to reflect packing, diameter and/or myelination of axons in white and gray matter.^29,30^

Mechanical forces generated by the rapid acceleration and deceleration of the brain during a concussion have a diffuse impact on brain tissues. In particular, stretching and shearing of axons is followed by progressive processes that result in chronic inflammation and conduction deficits,^31– 34^ with enhanced susceptibility at the gray-white matter interface.^35–37^ Unmyelinated axons in the developing brain may be especially vulnerable to disruption due to a lack of physical protection from myelin and exposure to a post-traumatic neuroinflammatory extracellular environment.^38^ The same diffusion MRI measures that are sensitive to the maturation of brain microstructure have been used to detect changes after concussion in the developing brain.^39,40^ Studies of pediatric concussion in the acute to post-acute phase (up to 1-month post-injury) demonstrate increased restricted diffusion (i.e. increased FA) suggesting increased axonal swelling post-injury.^41–46^ Whereas, studies of pediatric concussion in the chronic phase (3-12 months) post-injury have demonstrated reductions in restricted diffusion (i.e. decreased FA), suggestive of axon injury,^5,47^ or no significant differences post-injury.^45,48^ However, the long-term (>12 months), sex-specific effects of injury in childhood on brain microstructure have not, to our knowledge, been investigated. This is necessary for informing the impact of concussion on maturation of brain microstructure and associated cognitive performance.

Here we conducted an advanced diffusion MRI study to assess brain wide microstructure in children with a lifetime history of concussion. We used the ND measure from the restriction spectrum imaging (RSI) model of diffusion MRI, due to its sensitivity to the development of brain microstructure to 1) investigate sex-specific differences in white and gray matter microstructure in 9-to 10-year-old children with (*n* = 336) and without (*n* = 7368) a history of concussion, and 2) assess the association between altered microstructure and performance on attention and processing speed tests in children with concussion. ND was assessed in four tissue types: deep white matter, superficial white matter, subcortical structures, and cortex. We hypothesized that children with concussion would show differences in ND compared to children without concussion, and that ND in tissue types where differences were detected would be associated with lower scores on the attention and processing speed tests in children with concussion.

## Materials and methods

### Participants and demographic data

Participants in this study were part of the longitudinal Adolescent Brain Cognitive Development (ABCD) Study designed to investigate brain development and health. The ABCD Study collected data on physiological, psychological, and social measures, along with neuroimaging data from 11875 participants aged 9 to 10 years old. Participants were recruited from 21 different sites across the United States (US), and closely match the demographic characteristics of the US population (https://abcdstudy.org/; https://abcdstudy.org/families/procedures/). All sampling, recruitment, and assessment procedures for the ABCD Study are outlined in Garavan *et al*.^49^ All data used in this study is from the Curated Annual Release 2.0 (https://data-archive.nimh.nih.gov/abcd). Demographic data including age, sex (defined as sex assigned at birth), and pubertal status (based on a summary score of physical development) were collected from the Physical Health measure completed by parents, and total combined family income, highest parental education, and race and ethnicity were collected from the Parent Demographics Survey. History of medication taken in the two weeks before screening was collected from the Parent Medications Survey Inventory Modified from PhenX. Any missing demographic data were imputed using the R Multivariate Imputation by Chained Equations (mice) package to avoid deletion of participants.^50,51^ Siblings were randomly excluded from the sample to only include one sibling. Children with a history of concussion were compared to children with no history of brain injury.

### Concussion

Information on concussion history was collected from the Modified Ohio State University Traumatic Brain Injury Screen – Short Version.^52,53^ This is a parent-report of children’s lifetime history of head injuries with follow-up questions about the mechanism of injury, the age at injury, and whether the child experienced any loss of consciousness or a gap in their memory. For children with multiple head injuries, this information was recorded for all injuries. Children with concussion, defined as those that had reported a head injury with a loss of consciousness for less than 30 minutes or post-traumatic amnesia for less than 24 hours, were included in this study. This definition is consistent with a recently published study on the ABCD cohort that describes rates of attention-deficit/hyperactivity disorder (ADHD) in these children.^54^ Time since injury is defined as the time since the first injury when the participant experienced more than one concussion.

### Imaging measures

MRI, including T1-weighted and diffusion-weighted imaging, was collected for all participants across 21 sites, and on 28 different scanners, including Siemens Prisma, Philips, and GE 3T. The study’s MRI acquisitions, processing pipelines and quality control procedures are described in detail elsewhere^55,56^ and relevant details are summarized here. Participants were excluded when they had poor quality of brain scans, corrupted image files, and/or missing imaging data.

### Restriction spectrum imaging

Multi-shell diffusion-weighted images were acquired at six directions at b=500 s/mm2, 15 directions at b=1000 s/mm2, 15 directions at b=2000 s/mm2, and 60 directions at b=3000 s/mm2. ABCD provided pre-processed region-based measures compiled across subjects and summarized in a tabulated form.^56^ The RSI model was fit using a linear estimation approach as described in Hagler *et al*.^57^ The current study used mean ND values calculated for four categories of tissue: i) deep white matter, from 37 long range major white matter tracts labeled using AtlasTrack,^58^ ii) superficial white matter, sub-adjacent to 68 regions of the cortex labeled using anatomical parcellation from the Desikan-Killiany atlas,^59^ iii) 28 subcortical gray matter structures labeled using anatomic whole-brain segmentation,^60^ and iv) cortical gray matter in 68 regions of the cortex labeled with the Desikan-Killiany atlas.^59^

### Cognitive measures

Measures of cognition from the National Institute of Health (NIH) Toolbox were collected in participants in the ABCD study.^61^ In this study, brain measures were examined in relation to two cognitive tasks from the NIH Toolbox that were selected for their known sensitivity to concussion.^5,62^ The Flanker Inhibitory Control and Attention Task is an inhibition and conflict monitoring task that measures the ability to accurately respond to congruent versus incongruent contexts. Information processing speed was evaluated based on performance on the Pattern Comparison Processing Speed Task, which is a measure of rapid visual processing.^61,63^ The analysis presented in this study used age-corrected standard scores with a mean of 100 and a standard deviation of 15.^61^

### Statistical analyses

#### Comparisons of neurite density in children with and without concussion

All statistical analyses were performed using R.^51^ Linear mixed effects models were computed using the R lmerTest package, as demonstrated by the model formula below. Models were generated for mean ND values of each of the four tissue categories (deep white matter, superficial white matter, subcortical gray matter, and cortical gray matter). We investigated the relationship between group, sex, age, and their interactions. If three-way interaction terms had a *p*-value of more than 0.05, they were trimmed from the model and two-way interactions between group, sex, and age were examined. If two-way interactions had a *p*-value of more than 0.05, they were trimmed from the model and group, sex, and age were examined as main effects. Fixed effects covariates were puberty, total combined family income, and race and ethnicity, and scanner was included as a random effect. When between group interactions were significant they were assessed using the emmeans package in R.^51,64^

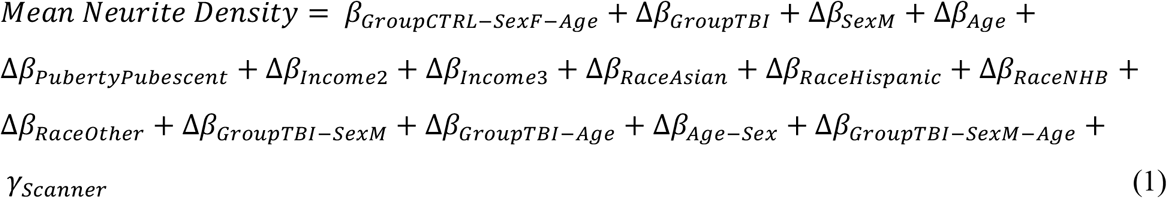

False discovery rate (FDR) correction was used to correct for Type 1 error rates across the four tissue categories (four comparisons: deep white matter, superficial white matter, subcortical structures, and cortex).^65^

##### Sensitivity analyses

Two sensitivity analyses were conducted in subsets of the full sample to assess the robustness of the group comparisons of ND. First, children with concussion were compared to an equally sized, closely matched comparison group. The comparison group was matched on age, sex, pubertal status, total combined family income, highest parental education, race and ethnicity, and medications taken in the last two weeks before screening including antipsychotics, antidepressants, and stimulants (based on a dichotomous response scale of yes/or no) and created using the R matchIt package.^51,66^ Second, the subset of children scanned on Siemens Prisma scanners, the most common scanner model, were examined. Linear mixed effect models as described above were generated to compare mean ND values in the concussion and comparison groups for each tissue category where differences were detected in the primary analyses.

#### Association between neurite density and injury variables in children with concussion

We investigated the relationship between injury characteristics and mean ND using linear mixed effects models, for tissue categories where significant differences were detected between children with and without concussion in the primary group comparison. Sex stratified analyses were conducted to investigate any differences in female and male children with concussion. Injury characteristics included injury mechanism, number of injuries, number of injuries with loss of consciousness, number of injuries with post-traumatic amnesia, age at injury, and time since first injury. FDR correction was used to correct for Type 1 error rates from multiple comparisons across analyses of each tissue category (two comparisons: deep white matter and superficial white matter).^65^

#### Association between neurite density and cognitive outcome in children with concussion

In the concussion group, dimensionality reduction of ND values of deep and superficial white matter regions was performed using principal component analysis (PCA) using the R psych package.^51^ Principal components (PCs) with eigenvalues (λ) greater than one were retained to extract a collection of regions that explain the largest variance in the data, indicated by the Kaiser-Meyer-Olkin (KMO) measure.^67^ Varimax rotation was applied to retained PCs so that each variable was associated with only one component and each component represented a small number of variables.^68^

Linear regression models were computed using the R lmerTest package.^51^ Models were generated to examine the association between each principal component for tissue categories where significant differences were detected between children with and without concussion, and age-corrected cognitive outcome scores, as demonstrated below. We investigated the relationship between PCs and sex, and their interaction. Significant interactions were followed by sex-stratified analyses to investigate the main effect of PCs on cognitive outcome scores within each sex separately. If there were no significant two-way interactions, the interaction term was trimmed from the model and the model was rerun to examine the main effect of PCs on cognitive outcome scores, with sex as a covariate.

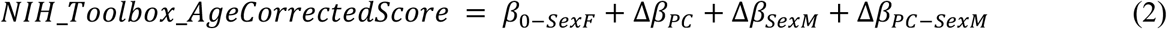

FDR correction was used to correct for Type 1 error rates from the comparisons of multiple PCs within analyses of each cognitive test and tissue category (10 comparisons for deep white matter and 18 comparisons for superficial white matter).^65^

### Data availability

All data used in this study is from the ABCD Study’s Curated Annual Release 2.0 (https://data-archive.nimh.nih.gov/abcd). Requests to access the derived data supporting the findings in this study should be directed to Anne Wheeler.

## Results

### Participants

This study examined children with concussion (*n* = 336) and children with no history of concussion (*n* = 7368) who had structural and diffusion images that passed quality control. The demographic characteristics of the two groups are presented in **Table 1** and the injury characteristics for the concussion group are presented in **Table 2**. For participants with more than one concussion, injury characteristics for all injuries are recorded. Age in months for the concussion group was statistically higher than the comparison group (*p* = .002), the concussion group had a greater number of male participants (*p* = .003), a greater number of participants from higher income households (*p* = .003), and a greater number of participants that were non-Hispanic white and fewer participants that were non-Hispanic black, Hispanic, Asian, or other/multi-racial than the comparison group (*p* < .001) (**Table 1**). The demographic characteristics of the matched comparison group and Siemens Prisma subsample are presented in **Supplementary Tables 1 and 2**.

**Table 1.** Demographic and characteristics.

|  | Concussion<br>(n = 336) |  | Comparison Group<br>(n = 7368) |  | p |
| --- | --- | --- | --- | --- | --- |
|  | M | F | M | F |  |
| **n** | 203 (60.4%) | 133 (39.6%) | 3843 (52.2%) | 3524 (47.8%) | **.003** |
| **Age in Months** | 120 (7.50) | 121 (7.42) | 119 (7.46) | 119 (7.34) | **.002** |
| **Pubertal Status:** |  |  |  |  |  |
| Prepubescence<sup>a</sup> | 150 (73.9%) | 34 (25.6%) | 2691 (70.0%) | 1068 (30.3%) | **.179** |
| Pubescence<sup>b</sup> | 53 (26.1%) | 99 (74.4%) | 1153 (30.0%) | 2456 (69.7%) |  |
| **Family Income<sup>c</sup>:** |  |  |  |  |  |
| < 50K | 47 (23.2%) | 30 (22.6%) | 1179 (30.7%) | 1116 (31.7%) | **.003** |
| 50-99K | 65 (32.0%) | 33 (24.8%) | 1044 (27.2%) | 1022 (29.0%) |  |
| > 100K | 91 (44.8%) | 70 (52.6%) | 1621 (42.2%) | 1386 (39.3%) |  |
| **Race:** |  |  |  |  |  |
| Non-Hispanic White | 124 (61.1%) | 97 (72.9%) | 2098 (54.6%) | 1817 (51.6%) | **<.001** |
| Non-Hispanic Black | 17 (8.4%) | 10 (7.5%) | 491 (12.8%) | 479 (13.6) |  |
| Hispanic | 35 (17.2%) | 16 (12.0%) | 806 (21.0%) | 770 (21.9%) |  |
| Asian<sup>d</sup> | 4 (2.0%) | 0 (0.0%) | 85 (2.2%) | 92 (2.6%) |  |
| Other/Multi-Racial | 23 (11.3%) | 10 (7.5%) | 364 (9.5%) | 366 (10.4%) |  |

<sup>a</sup>Prepubescence includes children at pubertal stages 1 and 2, as defined by the Physical Health Measure completed by Parents.
<sup>b</sup>Pubescence includes children at pubertal stages 3-5, as defined by the Physical Health Measure completed by Parents.
<sup>c</sup>Family Income refers to total combined family income.
<sup>d</sup>Asian race refers to Asian Indian, Chinese, Filipino, Japanese, Korean, Vietnamese, other Asian.
M, males; F, females.
P values were calculated using Independent t-test for continuous variables and chi-square test for categorical variables (p < .05 is bolded).

**Table 2.** Injury characteristics of concussion group.

|  | Concussion<br>(n = 336) |  | p |
| --- | --- | --- | --- |
|  | M | F |  |
| **n** | 203 (60.4%) | 133 (39.6%) | **<.001** |
| **Age at First Injury in Months** | 77.5 (31.5) | 69.5 (35.2) | .243 |
| **Time Since First Injury in Months** | 43.6 (31.1) | 54 (34.5) | .226 |
| **Injury Mechanism** |  |  |  |
| Play/Sports | 124 (61.1%) | 82 (61.7%) | **.998** |
| Motor Vehicle Accident | 9 (4.4%) | 6 (4.5%) |  |
| Fight/Shot/Shaken | 5 (2.5%) | 4 (3.0%) |  |
| ED Visit/Unknown Mechanism<sup>a</sup> | 55 (27.1%) | 35 (26.3%) |  |
| Other<sup>b</sup> | 10 (4.9%) | 6 (4.5%) |  |
| **Concussion Characteristics (injuries)** |  |  |  |
| With Loss of Consciousness <30 minutes | 43 (21.2%) | 36 (27.1%) | **.486** |
| With Post-Traumatic Amnesia <24 hours | 177 (87.2%) | 118 (88.7%) | **.324** |
| **Number of Concussions** |  |  |  |
| 1 | 174 (85.7%) | 117 (88.0%) | **.755** |
| 2 | 27 (13.3%) | 12 (9.0%) |  |
| 3 | 2 (1.0%) | 4 (3.0%) |  |

<sup>a</sup>Unknown mechanism of head injury that resulted in visit to the emergency department.
<sup>b</sup>Any other head injuries that lead to loss of consciousness or post-traumatic amnesia.
P values were calculated using chi-square test for categorical variables (p < .05 is bolded).
ED, emergency department; M, males; F, females.

### Comparisons of neurite density in children with and without concussion

#### White matter

When examining mean ND in both deep and superficial white matter, three-way interactions between group, sex, and age had a *p*-value of more than .05 and were trimmed from the final model. Age showed a robust positive relationship with mean ND of deep (β = 0.0004, *p* = <.001) and superficial white matter (β = 0.0005, *p* = <.001) (**Fig. 1A, Fig. 1B, Table 3**). Main effect of sex showed a significant relationship with mean ND of superficial white matter with lower ND in males than females (β = -0.003, *p* = <.001) (**Table 3**). Two-way interactions between group and sex were significant in deep white matter (β = -0.006, *p* = .013) and superficial white matter (β = -0.009, *p* = .004), with higher ND in female children with concussion, compared to the females without concussion (deep white matter: β = 0.004, *p* = .027; superficial white matter: β = 0.007, *p* = .001) (**Fig. 1C, Fig. 1D, Table 3**). There were no significant differences between males with and without concussion (deep white matter: β = -0.002, *p* = .118; superficial white matter: β = -0.002, *p* = .200).

**Figure 1.**
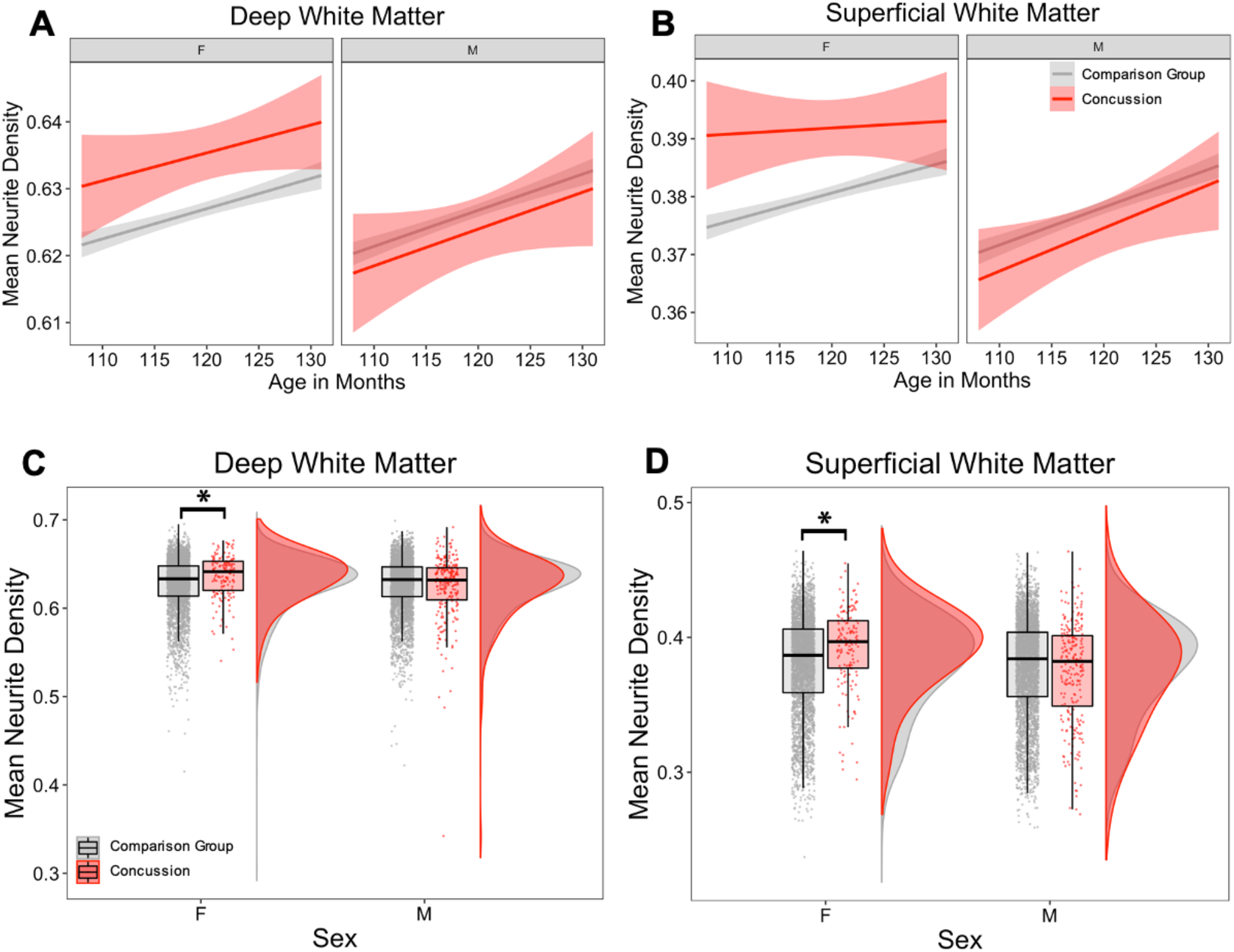
Group differences in mean neurite density between concussion and comparison groups, and sex. **(A-B)** Regression lines showing relationship between age and mean ND in children with concussion compared to the comparison group. **(A)** In the deep white matter and **(B)** superficial white matter, females with concussion had higher mean ND than comparison group, and males with concussion had lower mean ND than the comparison group, with similar trajectories with increasing age. **(C-D)** Mean ND in children with concussion and comparison group. Females with concussion had higher mean ND in (**C**) deep white matter and (**D**) superficial white matter. There were no group differences in males. Asterisks indicate significant group differences (*p* < .05).

**Table 3.** Group comparisons of mean neurite density between children with concussion and comparison group.

|  | Interaction |  |  |  |  |  | Main Effect |  |  |  |  |  | R<sup>2</sup> |
| --- | --- | --- | --- | --- | --- | --- | --- | --- | --- | --- | --- | --- | --- |
|  | Group by Sex |  | Female Sex |  | Male Sex |  | Group |  | Sex |  | Age |  |  |
|  | β | p | β | p | β | p | β | p | β | p | β | p |  |
| Cortical Gray Matter | - | - | - | - | - | - | -0.0002 | .817 | 0.0001 | .849 | **0.0002** | **<.001** | 0.500 |
| Subcortical Gray Matter | - | - | - | - | - | - | 0.0006 | .492 | -0.0003 | .653 | **0.0002** | **<.001** | 0.666 |
| Superficial White Matter | -0.009 | .004 | **0.007** | **.001** | -0.002 | .200 | - | - | **-0.003** | **<.001** | **0.0005** | **<.001** | 0.488 |
| Deep White Matter | **-0.006** | **.013** | **0.004** | **.027** | -0.002 | .118 | - | - | -0.001 | .178 | **0.0004** | **<.001** | 0.455 |

Corrected $p$ values based on FDR correction performed within analyses are reported and specified as significant ( $p < .05$ is bolded), with comparison group as the reference group.
Main effects of sex and age are reported for all models.
Main effects for group are reported for models where no interaction terms were retained.
R<sup>2</sup> represents conditional R<sup>2</sup> which takes both the fixed and random effects into account.

#### Gray matter

When examining mean ND in subcortical and cortical gray matter regions, the three-way and two-way interactions between group, sex, and age had a *p*-value of more than .05 and were trimmed from the final model. Age showed a robust positive relationship with mean ND (**Table 3**). There were no significant differences in mean ND between children with and without concussion in subcortical and cortical gray matter and no significant effects of sex (**Table 3**).

#### Sensitivity analyses

Sensitivity analyses of group comparisons of mean ND showed a similar pattern of results (**Supplementary Table 3**). Females with concussion showed higher mean ND in superficial white matter when compared to a closely matched comparison group (**Supplementary Fig. 1**), as well as when only the subset of children scanned on the Siemens Prisma scanners were examined (**Supplementary Fig. 2**). Comparisons of mean ND in deep white matter similarly revealed higher ND in females with concussion, however these comparisons did not reach statistical significance.

### Association between neurite density and injury variables in children with concussion

Children with a concussion from a motor vehicle crash showed lower ND in deep white matter (β = -0.018, *p* = .039). Sex-stratified analyses demonstrated that injury from play or sports was associated with higher ND of superficial white matter in males (β = 0.010, *p* = .049), but not in females. All results are reported in **Supplementary Table 4**.

### Association between neurite density and cognitive outcome in children with concussion

#### Deep white matter

PCA on ND of deep white matter regions generated four PCs with λ > 1. All PC loadings are presented in **Table 4**.

**Table 4.** Association between altered neurite density in deep white matter in concussion group and age-corrected scores on the Pattern Comparison Processing Speed Test.

|  | Interaction |  | Main Effect |  | R<sup>2</sup> | Loadings |
| --- | --- | --- | --- | --- | --- | --- |
|  | PC by Sex |  | PC |  |  |  |
|  | β | p | β | p |  |  |
| PC1 | - | - | -0.877 | 1.00 | 0.029 | R/L cingulate cingulum;<br>L anterior thalamic radiation<br>R uncinate<br>R/L inferior longitudinal fasciculus<br>R/L inferior frontooccipital fasciculus<br>Forceps major and minor<br>Corpus callosum<br>R/L temporal superior longitudinal fasciculus<br>R/L parietal superior longitudinal fasciculus<br>R/L striatal inferior frontal cortex<br>R/L inferior frontal superior frontal cortex |
| PC2 | - | - | 3.095 | .065 | 0.048 | R/L corticospinal/pyramidal tract<br>R anterior thalamic radiation<br>R/L superior corticostriate |
| PC3 | - | - | 0.436 | 1.00 | 0.028 | R/L parahippocampal cingulum<br>L uncinate |
| PC4 | - | - | 0.040 | 1.00 | 0.004 | R/L Fornix |

Corrected $p$ values are reported and bolded if $p < .05$ and italicized if $p < .1$ based on FDR correction performed within analyses.
R<sup>2</sup> represents adjusted R<sup>2</sup>.
Loadings represent regions of interest loaded on each principal component.
PC, principal component; R, right; L, left.

##### Flanker Inhibitory Control and Attention Test

No two-way interactions of PCs and sex, or main effects of PCs or sex were significantly associated with age-corrected scores of the Flanker Inhibitory Control and Attention Test.

##### Pattern Comparison Processing Speed Test

No two-way interactions of PCs and sex, or main effects of PCs and sex were significantly associated with age-corrected scores of the Pattern Comparison Processing Speed Test after correcting for multiple comparisons (**Table 4**).

#### Superficial white matter

PCA on ND of superficial white matter regions generated nine PCs with λ greater than one (average measure of sampling adequacy was 0.97). All PC loadings are presented in **Table 5**.

**Table 5.** Association between altered neurite density in superficial white matter in concussion group and age-corrected scores on the Pattern Comparison Processing Speed Test.

|  | Interaction |  | Main Effect |  | R<sup>2</sup> | Loadings |
| --- | --- | --- | --- | --- | --- | --- |
|  | PC by Sex |  | PC |  |  |  |
|  | β | p | β | p |  |  |
| PC1 | 6.273 | .115 | - | - | 0.045 | R/L superior temporal sulcus<br>R/L post- and pre- central gyri<br>R paracentral lobule<br>R/L frontal gyrus<br>R inferior frontal gyrus<br>R/L parietal lobe (superior, inferior)<br>R fusiform<br>R temporal gyrus<br>R cingulate (isthmus, posterior)<br>R precuneus<br>R lateral occipital |
| PC2 | - | - | 0.917 | 1.00 | 0.029 | R caudal anterior cingulate<br>L/R entorhinal cortex<br>R inferior temporal gyrus<br>L/R medial orbital frontal cortex<br>R parahippocampal<br>L/R rostral anterior cingulate<br>L/R temporal pole<br>R insula |
| PC3 | 8.410 | .008 | - | - | 0.063 | L temporal gyrus<br>R/L orbital frontal cortex<br>L parahippocampal gyrus<br>L inferior frontal gyrus<br>R pars orbitalis<br>L insula |
| PC4 | - | - | 0.399 | 1.00 | 0.028 | L caudal anterior cingulate<br>L transverse temporal |
| PC5 | - | - | 1.626 | .565 | 0.033 | R/L cuneus<br>L fusiform<br>L lateral occipital<br>L paracentral<br>R/L pericalcarine<br>R lingual |
| PC6 | - | - | -1.720 | .565 | 0.034 | L isthmus cingulate<br>L paracentral<br>L posterior cingulate<br>L precuneus |
| PC7 | - | - | 0.904 | 1.00 | 0.029 | R/L frontal pole |
| PC8 | - | - | 3.133 | .050 | 0.049 | No variables loaded onto this PC after varimax rotation |
| PC9 | - | - | 0.881 | 1.00 | 0.003 | No variables loaded onto this PC after varimax rotation |

Corrected *p* values are reported and bolded if *p* < .05 and italicized if *p* < .1 based on FDR correction performed within analyses.
R<sup>2</sup> represents adjusted R<sup>2</sup>.
Loadings represent regions of interest loaded on each principal component.
PC, principal component; R, right; L, left.

##### Flanker Inhibitory Control and Attention Test

No two-way interactions of PC and sex, or main effects of PCs or sex were significantly associated with age-corrected scores of the Flanker Inhibitory Control and Attention Test.

##### Pattern Comparison Processing Speed Test

A significant two-way interaction between PC3 and sex on Pattern Comparison Processing Speed Test performance was detected (β = 8.410, *p* = .008). This interaction was driven by a significant negative association in females in sex stratified analyses (β = -4.435, *p* = .024) indicating higher ND in these regions was associated with lower scores, and a significant positive association in males (β = 3.974, *p* = .004) indicating that higher ND in these regions was associated with higher scores (**Fig. 2A**). PC3 explained 11.36% of the variance and included positive loadings of fibers subjacent to the frontal and temporal lobes, including the left temporal gyrus, bilateral orbital frontal cortex, left parahippocampal gyrus, left inferior frontal gyrus, right pars orbitalis, and left insula **(Fig. 2B, Table 5)**.

**Figure 2.**
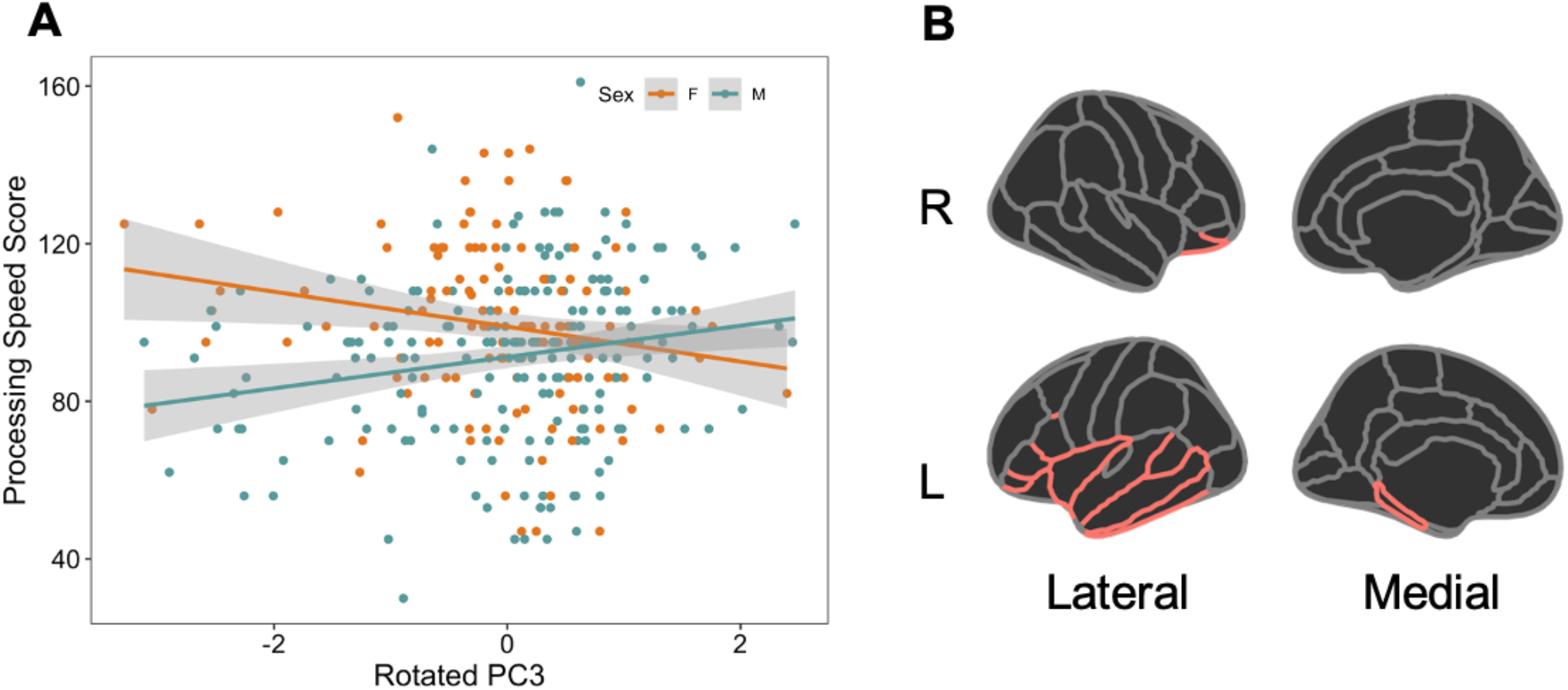
Relationship between mean neurite density and scores on the pattern comparison processing speed test. **(A)** Relationship between rotated PC3 of superficial white matter and age-corrected scores from the Pattern Comparison Processing Speed Test. Females with concussion showed poor performance of processing speed with increasing ND of regions that loaded onto PC3. **(B)** Desikan-Killiany atlas with red outline representing superficial white matter that loaded onto PC3. The red outline represents cortical regions from the Desikan-Killiany atlas adjacent to the superficial white matter that loaded on PC3. These regions include the left temporal, bilateral orbitalfrontal cortex, left parahippocampal gyrus, left inferior frontal gyrus, right pars orbitalis, left insula.

## Discussion

This study investigated cortical and white matter microstructure in 9-to 10-year-old children with and without concussion from the ABCD Study, using advanced diffusion MRI. ND derived from RSI showed a robust positive relationship with age across all brain tissue categories and increased ND was detected in superficial and deep white matter regions in females with a history of concussion compared to females without a history of concussion. When examining the relationships between altered microstructure and cognitive performance in children with concussion, higher ND in superficial white matter in frontal and temporal lobes was associated with lower scores on the processing speed test in females. These findings suggest that childhood concussion may lead to premature maturation of white matter in females and this altered trajectory may influence performance on cognitive tests. The current study is the first to use advanced multi-compartmental diffusion MRI modelling in a large sample of similarly aged children with concussion, allowing for the brain-wide investigation of altered microstructure.

Expected differences in brain microstructure related to age and sex were detected. Despite the narrow age range of 9 to 10 years old included in this study, small but robust positive relationships between age and ND were observed in white matter and gray matter. White matter and gray matter microstructural changes are evident in typical brain development.^69,70^ Prior diffusion MRI studies have demonstrated that throughout childhood, there is a steady increase in directionally restricted diffusion in white matter measured as FA from the commonly used simpler DTI model^24^ and ND from another advanced multicompartment model, neurite orientation dispersion and density imaging (NODDI).^71^ ND from both the NODDI model and the RSI model used in this study is more specific to brain microstructure than FA as it reflects intracellular directional diffusion which in the developing white matter is likely due to increased axon diameter, more densely packed axons, and/or increasing myelination leading to reduced water exchange.^29,72,73^ Literature investigating age-related changes in cortical gray matter report initial cortical thickening followed by cortical thinning of frontotemporal regions with age,^74–76^ which may be due to increasing synaptic pruning^75,77^ as well as age-related increases in myelin content.^78^ As pruning and associated reductions in dendritic arborization would be expected to decrease ND, the age-related increases in ND observed in cortical and subcortical gray matter in this study are likely driven by this increased myelin content.^29^ The current study also detected a main effect of sex in superficial white matter indicating that females have higher ND than males, consistent with reports that female children show earlier diffusion changes compared to males.^24,79^

When comparing brain microstructure in children with and without concussion, differences were detected in superficial and deep white matter regions, but not in cortical or subcortical gray matter. These findings support the vulnerability of white matter to chronic effects of concussion,^33,80^ and support current literature that reports differences relative to controls, and changes over time in diffusion measures in white matter after concussion in childhood.^36,81–83^ Moreover, we observed stronger and more robust differences in superficial white matter than in deep white matter as these remained significant for comparisons with both the closely matched comparison group as well as the subset of children scanned on the Siemens Prisma scanners in our sensitivity analyses. This may be due to the protracted time course of superficial white matter myelination making it more vulnerable to childhood injury.^24,84,85^ Damage to superficial white matter fibers has been reported in post-mortem studies of traumatic brain injury,^86,87^ and diffusion differences in superficial white matter related to concussion were recently described by our group in another large pediatric population cohort with a wider age range.^36^ The complex patterns of crossing fibers in these short tracts are likely more accurately captured in the current study due to the use of RSI over the simpler DTI model.^85^

In this study, the timing of injury relative to the detected white matter differences, and the age of the participants are critical aspects to consider when placing the results in the context of existing studies of white matter microstructure in children with concussion. The children in the current study were assessed an average of almost four years after concussion, thus the increase in ND is unlikely to reflect acute injury effects and the direction of effect is inconsistent with previous studies that described less restricted diffusion (i.e. decreased FA) at chronic timepoints.^5,45,47,48^ Unique aspects of the current study may explain the apparent discrepancy. First, the restricted age range of 9-to 10-year-olds is at the lower end of the age ranges included in previous studies which may have increased the sensitivity of the group comparison as it reduced developmental heterogeneity in the sample. Second, the large sample size in this study exceeds previous studies allowing for the detection of small effects. Third, the use of multi-compartment diffusion MRI modeling rather than the more common DTI model may allow for more sensitive and specific estimates of brain microstructure.^85^ Considering the existing literature, unique aspects of the current study, and strong association between ND and age, one possibility is that the observed effects reflect that concussion early in life accelerates the developmental maturation of white matter, rather than injury pathology in white matter per se. Thus, higher ND observed in female children with past concussion in the current study may reflect increased axon diameter, more densely packed axons and/or increasing myelination, reflecting a state of premature white matter maturation. Comparable profiles of atypical development of brain connectivity have been observed following pre- and post-natal adversity in other studies.^88,89^ The current study is the first, to our knowledge, to show sex specific differences in diffusion measures between children with and without concussion. Previous studies which investigated post-acute diffusion in white matter microstructure following concussion did not detect significant interaction effects between sex and concussion,^43,45^ suggesting that sex specific effects of concussion on white matter microstructure may only emerge once the physiological changes induced by concussion have had time to interact with neurodevelopmental processes.

When evaluating cognitive performance in children with concussion, significant associations between ND and scores on the processing speed task but not the attention task were detected. Previous studies have identified both attention and processing speed as cognitive domains that are particularly impaired after concussion in children,^5,6^ though impairments after a childhood concussion normalize in 70-90% of children 3 to 12 months post-injury.^11–13^ Previous studies that examined associations between diffusion measures in white matter and these cognitive domains have generally found that less restricted diffusion (i.e. decreased FA) is associated with poorer performance,^36,90^ consistent with the observation in this study in males where decreased ND in superficial white matter was associated with lower scores on the processing speed test. However, in the current study, *increased* ND was associated with lower scores on the processing speed test in superficial white matter in females. Future research may examine if the sex-specific direction of this relationship and the specificity of the observed differences in white matter microstructure in females provides insight into the neurobiological mechanisms of increased prevalence of persistent symptoms in female children with concussion.^14,91^

Some aspects of the study sample and methodology limit the conclusions that can be made from these analyses. First, the children with concussion experienced their injury at a wide range of ages, which may have differential effects on brain development. However, an effect of age at injury on white matter microstructure was not detected when assessing the impact of injury variables on brain microstructure. Second, despite the inclusion of possible confounding factors as covariates in the statistical models and sensitivity analysis in a closely matched comparison sample, the cross-sectional design of this study means that we cannot rule out that differences in brain white matter are associated with a factor that predisposed females to brain injury rather than were a consequence of brain injury. Third, due to the heterogeneity of brain regions impacted by concussion, we did not hypothesize region-specific effects of injury and instead examined whole brain effects of mean ND across four tissue categories (deep white matter, superficial white matter, subcortical gray matter, and cortical gray matter). This may have obscured any effects specific to vulnerable brain regions and may have contributed to the small effect sizes observed in our analyses. Furthermore, although RSI allows for the distinction between intracellular and extracellular water diffusion and provides more specific information than DTI, the exact microstructural drivers of the observed differences in ND are not known.

## Conclusion

This study identified increased ND in white matter of female children aged 9-10 with a history of concussion. These differences may reflect premature maturation of white matter with a possible impact on processing speed. This study is the first to use multi-compartmental diffusion MRI to investigate the sex-specific effects of childhood concussion on white matter microstructure and lays the foundation for future analyses that track the maturation of white matter in these children.

## Data Availability

Requests to access the derived data supporting the findings in this study should be directed to Anne Wheeler.

https://data-archive.nimh.nih.gov/abcd

## Abbreviations

ABCD: Adolescent Brain Cognitive Development
ATR: Anterior Thalamic Radiation
DTI: Diffusion Tensor Imaging
FA: Fractional Anisotropy
FDR: False Discovery Rate
IFG: Inferior Frontal Gyrus
IFOF: Inferior Fronto-occipital Fasciculus
ILF: Inferior Longitudinal Fasciculus
ND: Neurite Density
NIH: National Institute of Health
NODDI: Neurite Orientation Dispersion and Density Imaging
OFC: Orbital Frontal Cortex
PC: Principal Component
RSI: Restriction Spectrum Imaging
SLF: Superior Longitudinal Fasciculus;

## Supplementary material

Supplementary material is available at *Brain* online.

