## Supplementary Tables and Figures for "Premature white matter microstructure in female children with a history of concussion"

**Supplementary Table 1 Demographic characteristics of the matched comparison group**

| | Concussion (*n* = 336) | | Matched Comparison Group (*n* = 336) | | *p* |
|---|---|---|---|---|---|
| | M | F | M | F | |
| *n* | 203 (60.4%) | 133 (39.6%) | 199 (61.3%) | 130 (38.7%) | .813 |
| **Average Age in Months** | 120 (7.50) | 121 (7.42) | 119 (7.38) | 120 (7.59) | .167 |
| **Pubertal Status** | | | | | |
| Prepubescence[a] | 150 (73.9%) | 34 (25.6%) | 146 (70.9%) | 35 (26.9%) | .816 |
| Pubescence[b] | 53 (26.1%) | 99 (74.4%) | 60 (29.1%) | 95 (73.1%) | |
| **Family Income[c]** | | | | | |
| < 50K | 47 (23.2%) | 30 (22.6%) | 49 (23.8%) | 24 (18.5%) | |
| 50-99K | 65 (32.0%) | 33 (24.8%) | 58 (28.2%) | 38 (29.2%) | .888 |
| > 100K | 91 (44.8%) | 70 (52.6%) | 99 (48.1%) | 68 (52.3%) | |
| **Race** | | | | | |
| Non-Hispanic White | 124 (61.1%) | 97 (72.9) | 118 (57.3%) | 84 (64.6%) | |
| Non-Hispanic Black | 17 (8.4%) | 10 (7.5%) | 32 (15.5%) | 10 (7.7%) | |
| Hispanic | 35 (17.2%) | 16 (12.0%) | 38 (18.4%) | 23 (17.7%) | .203 |
| Asian[d] | 4 (2.0%) | 0 (0.0%) | 4 (1.9%) | 1 (0.8%) | |
| Other/Multi-Racial | 23 (11.3%) | 10 (7.5%) | 14 (6.8%) | 12 (9.2%) | |

*P* values were calculated using Independent t-test for continuous variables and chi-square test for categorical variables.
[a]Prepubescence includes children at pubertal stages 1 and 2, as defined by the Physical Health Measure completed by Parents.
[b]Pubescence includes children at pubertal stages 3-5, as defined by the Physical Health Measure completed by Parents.
[c]Family Income refers to total combined family income.
[d]Asian race refers to Asian Indian, Chinese, Filipino, Japanese, Korean, Vietnamese, other Asian.
M, males; F, females.

**Supplementary Table 2 Demographic and injury characteristics for participants scanned on Siemens Prisma scanners**

| | Concussion Scanned on Siemens (*n* = 223) | | Siemens Comparison Group (*n* = 4498) | | *p* |
|---|---|---|---|---|---|
| | M | F | M | F | |
| *n* | 128 (57.4%) | 95 (42.6%) | 2381 (52.9%) | 2117 (47.1%) | .192 |
| **Average Age in Months** | 121 (7.35) | 120 (7.56) | 119 (7.41) | 119 (7.32) | **.010** |
| **Pubertal Status** | | | | | |
| Prepubescence[a] | 94 (73.4%) | 23 (24.2%) | 1612 (67.7%) | 632 (29.9%) | .452 |
| Pubescence[b] | 34 (26.6%) | 72 (75.8%) | 769 (32.3%) | 1485 (70.1%) | |
| **Family Income[c]** | | | | | |
| < 50K | 32 (25.0%) | 21 (22.1%) | 715 (30.0%) | 630 (29.8%) | |
| 50-99K | 43 (33.6%) | 24 (25.3%) | 648 (27.2%) | 672 (31.7%) | .118 |
| > 100K | 53 (41.4%) | 50 (52.6%) | 1018 (42.8%) | 815 (38.5%) | |
| **Race** | | | | | |
| Non-Hispanic White | 82 (64.1%) | 74 (77.9%) | 1336 (56.1%) | 1154 (54.5%) | |
| Non-Hispanic Black | 14 (10.9%) | 8 (8.4%) | 368 (15.5%) | 321 (15.2%) | |
| Hispanic | 20 (15.6%) | 9 (9.5%) | 431 (18.1%) | 417 (19.7%) | **<.001** |
| Asian[d] | 3 (2.3%) | 0 (0%) | 34 (1.4%) | 36 (1.7%) | |
| Other/Multi-Racial | 9 (7.0%) | 4 (4.2%) | 212 (8.9%) | 189 (8.9%) | |

*P* values were calculated using Independent t-test for continuous variables and chi-square test for categorical variables.
[a]Prepubescence includes children at pubertal stages 1 and 2, and pubescence includes children at pubertal stages 3-5, as defined by the Physical
[b]Health Measure completed by Parents.
[c]Family Income refers to total combined family income.
[d]Asian race refers to Asian Indian, Chinese, Filipino, Japanese, Korean, Vietnamese, other Asian.
M, males; F, females.

**Supplementary Table 3 Group comparisons of mean neurite density between children with concussion and matched comparison group and Siemens Prisma group**

| | | Interaction | | | | Main Effect | | | | |
|---|---|---|---|---|---|---|---|---|---|---|
| | | Group by Sex | | Age by Sex | | Female Sex | | Age | | |
| | | β | *p* | β | *p* | β | *p* | β | *p* | $R^2$ |
| **Matched Comparison** | **SWM** | **-0.009** | **.039** | - | - | **0.007** | **.041** | **0.0007** | **<.001** | 0.498 |
| | **DWM** | *-0.007* | *.070* | - | - | 0.005 | .121 | **0.0007** | **<.001** | 0.470 |
| **Siemens Prisma** | **SWM** | **-0.010** | **.007** | - | - | **0.007** | **.008** | **0.0005** | **<.001** | 0.182 |
| | **DWM** | **-0.006** | **.046** | **0.0002** | **.040** | 0.003 | .133 | **0.0004** | **<.001** | 0.089 |

*P* < .05 is bolded, *p* < .1 is italicized with comparison group as the reference group.
Main effects of sex and age are reported for all models.
$R^2$ represents conditional $R^2$ which takes both the fixed and random effects into account.
SWM, superficial white matter; DWM, deep white matter.

**Supplementary Table 4 Association between neurite density in the concussion group and injury variables**

| | Superficial White Matter | | | | | | Deep White Matter | | | | | |
|---|---|---|---|---|---|---|---|---|---|---|---|---|
| | All | | Female | | Male | | All | | Female | | Male | |
| | β | *p* | β | *p* | Β | *p* | β | *p* | β | *p* | β | *p* |
| **Injury Mechanism** | | | | | | | | | | | | |
| Fight | 0.005 | .637 | 0.004 | .760 | 0.016 | .302 | 0.006 | .637 | 0.012 | .338 | 0.009 | .577 |
| MVA | -0.0005 | .945 | -0.013 | .299 | 0.008 | .479 | **-0.018** | **.039** | -0.015 | .172 | *-0.019* | *.097* |
| Play/Sports | 0.003 | .890 | -0.004 | .449 | **0.010** | **.049** | 0.0001 | .979 | -0.004 | .396 | 0.003 | .596 |
| Other | -0.002 | .806 | -0.005 | .726 | 0.003 | .832 | 0.004 | .806 | -0.003 | .809 | 0.008 | .603 |
| **Number of Injuries** | 0.0002 | .969 | 0.003 | .647 | -0.011 | .235 | -0.002 | .969 | -0.002 | .773 | -0.008 | .364 |
| w/ Loss of Consciousness <30 minutes | 0.004 | .580 | *0.010* | *.066* | 0.002 | .710 | -0.002 | .667 | 0.008 | .109 | -0.007 | .276 |
| w/ Post-Traumatic Amnesia <24 hours | 0.006 | .477 | -0.001 | .904 | *0.013* | *.064* | 0.003 | .505 | -0.0004 | .953 | 0.006 | .437 |
| **Age at First Injury** | -0.00002 | .912 | -0.0001 | .668 | 0.0002 | .544 | -0.00003 | .912 | -0.00002 | .939 | 0.0001 | .687 |
| **Time Since First Injury** | -0.00001 | .946 | -0.0002 | .366 | 0.0003 | .402 | 0.00002 | .946 | -0.0001 | .612 | 0.0002 | .459 |

*P* < .05 is bolded, *p* < .1 is italicized.
Effects are reported for concussion group analyses and sex stratified analyses.

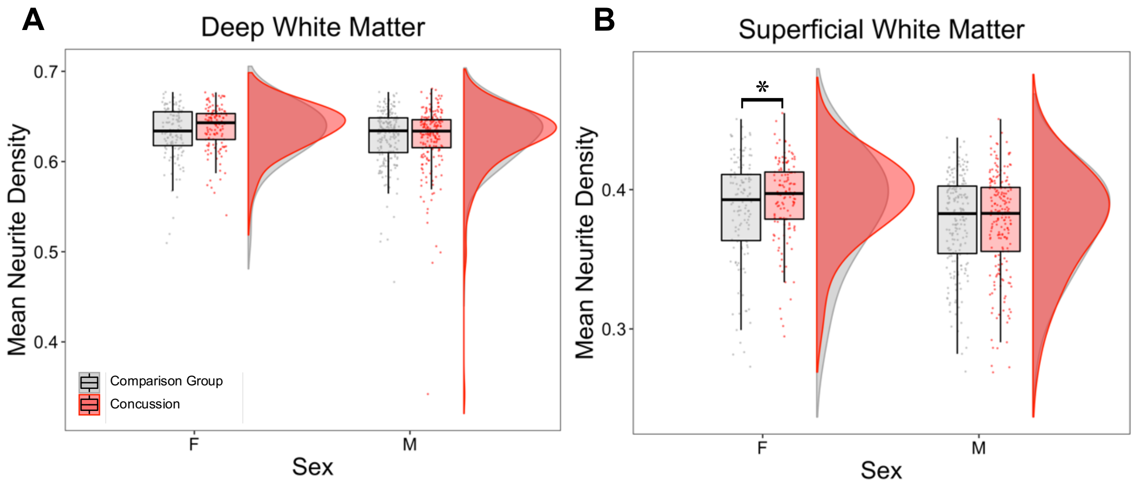


**Supplementary Figure 1 Mean ND in children with concussion compared to matched comparison group. (A)** Deep white matter. **(B)** Superficial white matter.

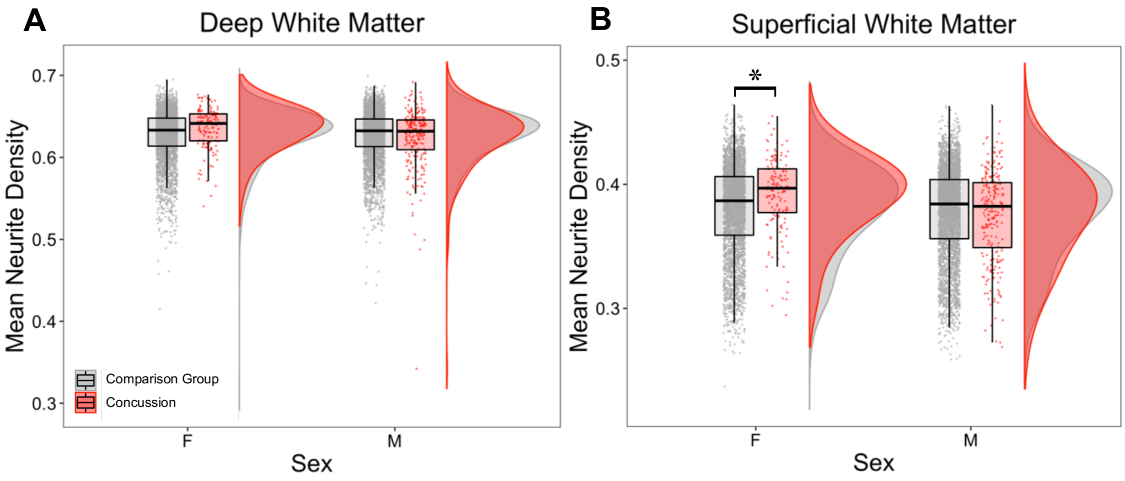


**Supplementary Figure 2 Mean ND from Siemens scanners in children with concussion compared to comparison group**. **(A)** Deep white matter. **(B)** Superficial white matter.
